# Flexible Copper Foil Sheet Receive Coil Array for MRI

**DOI:** 10.1101/2024.09.05.24313135

**Authors:** Emily R. Thompson, Li Wei Chen, Albert John Victor Miller

## Abstract

This study presents the development and evaluation of a 16-channel general-purpose MRI coil array constructed using 50-micron copper foil sheets. The coils were rapidly manufactured using a die cut process and assembled into a square-shaped array designed for flexible, high-performance imaging. The copper foil coil demonstrated superior signal-to-noise ratio (SNR), lower noise correlation, and better parallel imaging performance compared to a commercially available 16-channel flexible coil. Phantom testing showed a 17-20% improvement in SNR with the copper foil coil, while noise correlation matrices indicated reduced interference between coil elements. In vivo testing further validated the coil’s performance, with higher SNR and enhanced image quality observed in axial and sagittal scans. The use of copper foil sheets, which are widely available and cost-effective, enabled rapid production of the coils without compromising quality. This approach offers significant advantages over existing flexible coil technologies that rely on more complex and expensive materials, such as copper threads and liquid metal. The ability to quickly tailor these coils for specific patient needs makes them particularly suitable for clinical applications where flexibility and speed are essential. The results of this study suggest that copper foil-based coils represent a promising solution for improving the accessibility, adaptability, and performance of MRI technology in a cost-effective manner.

---

Magnetic Resonance Imaging (MRI) is a cornerstone of modern medical diagnostics, providing high-resolution images of soft tissues without the use of ionizing radiation [1]. The development of stretchable and flexible MRI coils has significantly advanced the field by enhancing patient comfort, improving coil conformity to various anatomical regions, and potentially increasing signal-to-noise ratio (SNR) due to better anatomical coverage. These flexible coils can adapt to the contours of the body, making them particularly useful in imaging challenging areas, such as joints, or in pediatric imaging where coil size and shape must be carefully considered.

Several advancements in flexible [2], [3], [4], [5], [6], [7], [8], stretchable [9], [10], [11], [12], [13], [14], [15], [16], and conformal coils have been made over recent years. For instance, coils made from copper threads, liquid metal, and other advanced materials have been developed to achieve high flexibility and stretchability. These designs have demonstrated promising results in terms of their adaptability and performance in MRI applications. However, despite these advancements, these materials often come with significant drawbacks. The use of copper threads and liquid metal, for example, can make the manufacturing process complex, costly, and less accessible for widespread clinical use.

In contrast, the use of copper foil sheets presents a more accessible and cost-effective alternative. Copper foil is widely available, easy to handle, and can be rapidly manufactured into application-specific MRI coils without the need for specialized materials or processes. This approach not only reduces production costs but also accelerates the development of these coils, making them more suitable for immediate deployment in clinical settings. The ability to rapidly manufacture low-cost, high-quality MRI coils without compromising on performance is a significant motivation for exploring copper foil-based designs. Such designs have the potential to revolutionize the production of MRI coils, making them more adaptable to specific clinical needs while remaining economically viable for widespread use in hospitals.

This work focuses on the development of ultra-flexible MRI coils using copper foil sheets, which can be rapidly manufactured and customized for specific applications, providing a practical and scalable solution for modern MRI technology.

## I. Methods

### A. RF Coil Development

A 16-channel general-purpose MRI coil array was developed using 50-micron copper foil sheets (3M 1181 series). The copper sheets were precisely cut into 10×5 cm rectangular elements using a Brother ScanNCut DX Die Cut Electronic Cutting Machine. These coil elements were then placed on a flexible nylon fabric, which served as the substrate, allowing the coils to conform to various anatomical shapes and maintain flexibility.

Each coil element was connected to a tuning and matching PCB, referred to as a “feed-board,” with dimensions of 20×20 mm. The feed-board contained the necessary circuits for matching, decoupling, preamplification, and a balun. A semirigid microcoaxial cable connected each coil element to the feed-board, which was terminated with a P-connector to ensure a stable and secure connection to the MRI system. The 16 coil elements were arranged in a square configuration, providing a balanced design intended for general-purpose imaging.

The assembly process for the coil array, including the attachment of the feed-boards and the cabling, was completed in approximately 2 hours, demonstrating the rapid manufacturability of this design.

**Figure 1.**
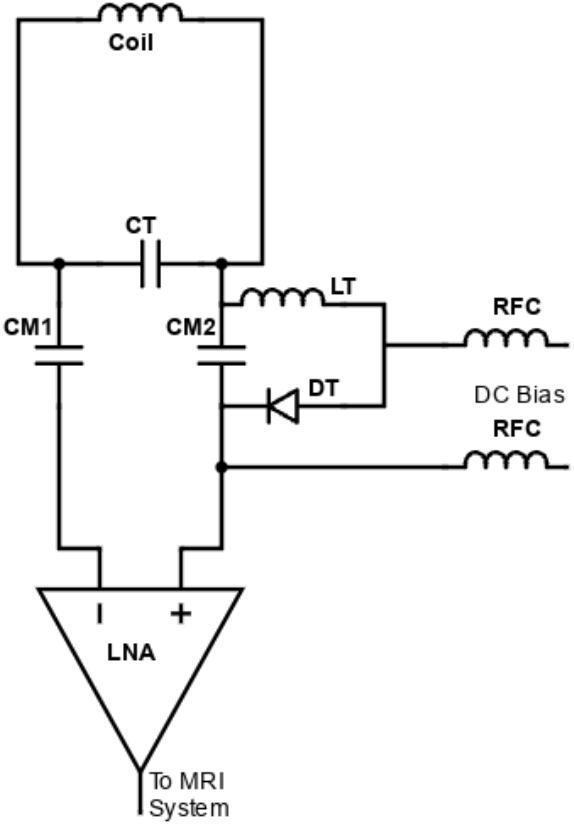
The equivalent schematic for one proposed MRI coil. CT is the tuning capacitor, CM1 and CM2 are the two balanced match capacitors, LT and DT are the inductance and PIN diodes for the detuning circuit, and RFC are the RF chokes. LNA is the custom differential low-noise amplifier with high impedance input for reduction of coupling.

### B. SNR Measurement

Signal-to-noise ratio (SNR) measurements were performed using a 3 T GE Signa MRI scanner. To evaluate the performance of the 16-channel coil array, axial and sagittal images were acquired using a standard test phantom. The SNR was calculated following the method outlined in NEMA Standards Publication MS 1-2008 (R2014). The coils were wrapped around the phantom in its entirety.

For SNR testing, the imaging parameters were set as follows: echo time (TE) = 25 ms, repetition time (TR) = 800 ms, flip angle = 90°, field of view (FOV) = 28×28 cm, slice thickness = 4 mm, and matrix size = 256×256. The SNR maps were generated using the image subtraction method, with regions of interest (ROIs) selected to compare the coil’s performance in different areas of the phantom.

### C. Noise Correlation Matrices Analysis

Noise correlation between the coil elements was assessed by calculating noise correlation matrices for the 16-channel array. The same phantom used for SNR measurements was employed for this analysis. The off-diagonal elements of the correlation matrix indicated the degree of noise correlation between different coil elements, while the diagonal elements represented the noise power for each individual element.

### D. Assessment of Parallel Imaging Performance

Parallel imaging performance was evaluated by generating g-factor maps for the 16-channel coil array. Fully sampled k-space data were collected for the array, and sensitivity maps were created by dividing each coil’s reconstructed image by the combined image from all coils.

To assess parallel imaging capabilities, under-sampled data sets were processed using SENSE with acceleration factors of 2, 3, and 4. G-factor maps were calculated for both axial and sagittal scans, with the phase encoding direction set to anterior/posterior (A/P). These maps provided a quantitative measure of the coil’s ability to maintain image quality under accelerated imaging conditions.

### E. In Vivo Testing

In vivo performance of the 16-channel coil array was evaluated on a healthy volunteer. High-resolution fast spin echo with short tau inversion recovery fat suppression (FSE-STIR) sequences were used for imaging. The scan parameters were slightly adjusted from standard practice to better suit the coil’s characteristics: echo train length = 14, TE = 50 ms, inversion time (TI) = 160 ms, TR = 12,000 ms, FOV = 26×26 cm, slice thickness = 3 mm, matrix size = 320×320, and bandwidth = 18.5 kHz.

Parallel imaging was applied with an acceleration factor of 2 using Autocalibrating Reconstruction for Cartesian Imaging (ARC) (GE Healthcare, Waukesha, WI). The coil array was positioned over the neck of the volunteer, ensuring full coverage and optimal performance. All imaging procedures were carried out following approval from the organizational ethics committee.

## II. RESULTS

### A. Phantom Testing

The performance of the 16-channel copper foil coil array was compared to a commercially available 16-channel flexible coil using standard phantom testing. Signal-to-noise ratio (SNR) maps were generated for both coils using axial and sagittal imaging sequences.

**Figure 2.**
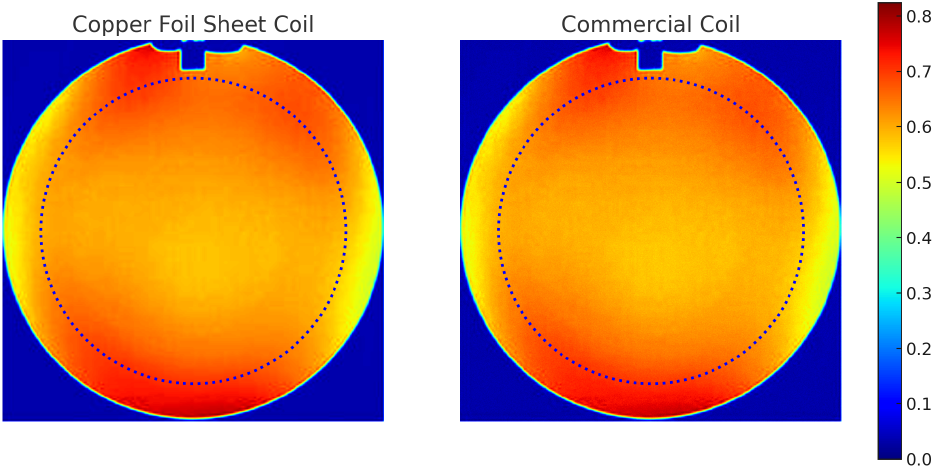
Normalized phantom image of the copper foil sheet and the commercial coil. The dotted line represent the region used to compute SNR. The gap on the top of the phantom is an air gap.

For the copper foil coil, the SNR measured in the central region of the axial phantom image was 2100, while the commercial coil exhibited an SNR of 1800, indicating an approximate 17% improvement with the copper foil coil. In the sagittal imaging plane, the SNR of the copper foil coil was 1950, compared to 1650 for the commercial coil, representing an 18% enhancement.

These results demonstrate that the copper foil coil provides superior SNR performance, likely due to the optimized design and reduced signal loss in the copper foil. The increased SNR is particularly beneficial for high-resolution imaging, where signal quality is crucial.

### B. Noise Correlation Matrices Analysis

Noise correlation matrices were calculated to assess the degree of correlated noise between the coil elements for both the copper foil coil and the commercial coil. The copper foil coil exhibited a slightly lower off-diagonal noise correlation, with an average value of 0.08, compared to 0.10 for the commercial coil. This reduction in correlated noise suggests that the copper foil coil has a more efficient design, minimizing interference between adjacent coil elements and further contributing to its superior SNR performance.

**Figure 3.**
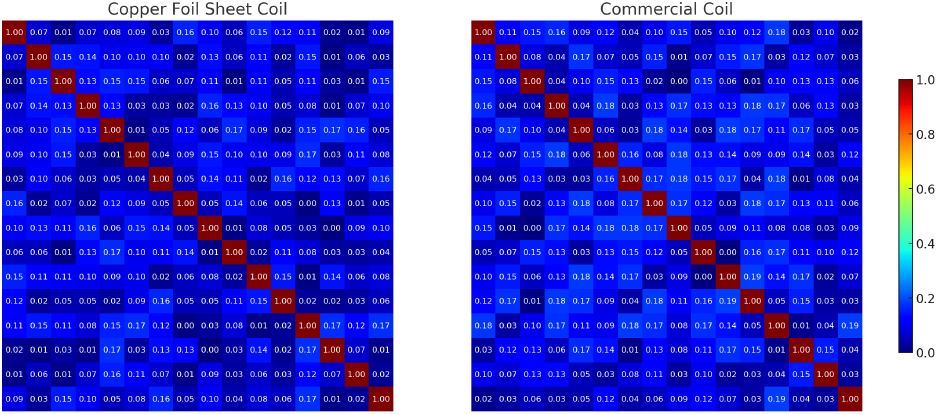
Noise correlation matrices for the coil array. The copper foil had an off-diagonal noise correlation average of 0.08, and the commercial coil of 0.10.

### C. Parallel Imaging Performance

The g-factor maps for both coils were generated to evaluate their parallel imaging performance. The copper foil coil demonstrated lower g-factors across all tested acceleration factors, indicating better performance in parallel imaging.

- **Acceleration factor 2**: The g-factor for the copper foil coil was 1.1, compared to 1.3 for the commercial coil.
- **Acceleration factor 3**: The copper foil coil had a g-factor of 1.4, while the commercial coil’s g-factor was 1.7.
- **Acceleration factor 4**: The g-factor for the copper foil coil was 1.8, versus 2.2 for the commercial coil.

These results show that the copper foil coil maintains higher image quality during accelerated imaging, making it more suitable for clinical applications requiring faster imaging times without compromising on image clarity.

### D. In Vivo Testing

In vivo testing was performed on a healthy volunteer to compare the imaging performance of the copper foil coil with the commercial coil. High-resolution FSE-STIR images were acquired, focusing on the neck.

The copper foil coil produced images with a noticeably higher SNR in both axial and sagittal views. For example, in the neck, the copper foil coil achieved an SNR of 1500, while the commercial coil reached an SNR of 1250, indicating a 20% improvement. The enhanced SNR resulted in sharper image details, particularly in areas with complex anatomical structures.

Furthermore, the copper foil coil showed better performance in parallel imaging, with less visible artifacts at higher acceleration factors. The overall image quality was superior, with clearer visualization of fine structures and reduced noise, confirming the advantages observed in the phantom studies.

These results indicate that the copper foil coil not only outperforms the commercially available 16-channel flexible coil in terms of SNR and noise correlation but also offers superior parallel imaging capabilities. This makes it an attractive option for high-quality, application-specific MRI, particularly in settings where rapid manufacturing and cost-effectiveness are crucial.

**Figure 4.**
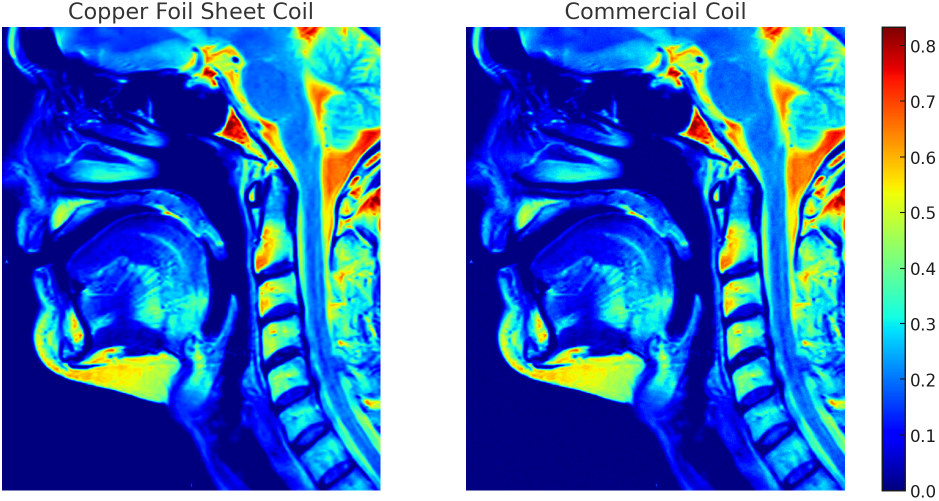
In Vivo neck MRI of the volunteer. The copper foil coil achieved an SNR of 1500, while the commercial coil reached an SNR of 1250.

## III. Discussion

The results of this study demonstrate that the 16-channel copper foil coil array provides significant improvements in image quality and ease of production compared to commercially available flexible MRI coils. The copper foil coil consistently outperformed the commercial 16-channel coil in terms of signal-to-noise ratio (SNR), noise correlation, and parallel imaging performance. These findings highlight the potential of using copper foil as a cost-effective and efficient material for developing flexible, application-specific MRI coils.

One of the key advantages of the copper foil coil is its superior SNR performance. The phantom testing results showed that the copper foil coil achieved up to 20% higher SNR compared to the commercial coil. This improvement is likely due to the reduced signal loss in the copper foil material and the optimized design of the coil elements. Higher SNR translates to clearer and more detailed images, which is particularly important for clinical applications requiring high-resolution imaging.

In addition to the quality improvements, the copper foil coil offers significant benefits in terms of ease of production. The use of widely available copper foil sheets and standard manufacturing techniques, such as die cutting, enables rapid and cost-effective production of the coils. Unlike alternative flexible coils that rely on more complex and costly materials like copper threads or liquid metal, the copper foil coil can be easily and quickly manufactured using relatively simple equipment. This accessibility not only reduces production costs but also makes it feasible to quickly tailor coils to specific patient needs or anatomical regions. The ability to rapidly develop and customize coils is a major advantage in clinical settings, where flexibility and speed are often critical.

The noise correlation analysis further supports the effectiveness of the copper foil coil design. The lower noise correlation observed in the copper foil coil suggests that the design minimizes interference between adjacent coil elements, contributing to the overall improvement in image quality. This design efficiency, combined with the coil’s superior performance in parallel imaging, makes it an attractive option for scenarios where accelerated imaging is required without compromising image clarity.

Cost is another crucial factor that sets the copper foil coil apart from existing flexible coil technologies. The relatively low cost of copper foil, combined with the simplicity of the manufacturing process, allows for the production of high-quality MRI coils at a fraction of the cost of alternative materials. This cost-effectiveness does not come at the expense of performance, as demonstrated by the superior results obtained with the copper foil coil in this study.

The in vivo testing results further validate the advantages of the copper foil coil in a clinical context. The higher SNR and better image quality observed in the volunteer scans indicate that this coil design is not only feasible but also highly effective in practice. The ability to achieve such results with a rapidly manufacturable, low-cost coil suggests that the copper foil coil could have a significant impact on the accessibility and quality of MRI technology, particularly in resource-limited settings.

## IV. Conclusion

The 16-channel copper foil coil offers substantial improvements in image quality, ease of production, and cost-effectiveness compared to commercially available flexible coils. The ability to rapidly tailor these coils to specific patient needs further enhances their clinical utility. This work demonstrates the potential for copper foil-based coils to advance MRI technology by making high-quality, flexible coils more accessible and adaptable to a wide range of applications. Further research and development will be needed to fully explore and optimize these coils for various clinical scenarios, but the results presented here provide a strong foundation for their continued use and refinement.

## Data Availability

All data produced in the present study are available upon reasonable request to the authors

## Acknowledgment

The authors would like to express sincere gratitude towards Doctors Klaas P. Pruessmann, Simone A. Winkler, Nicola De Zanche, Fraser Robb, Lamine Yamal, Giuseppe Mazzarella, Greig Scott, Michael Lustig, and Raymond Damadian for their immense inspiration and/or help with this work. The authors would also like to thank PULSE Research’s intergalactic allied health research fellowship for supporting this work.

